# Impact of PET reconstruction on Aβ-amyloid quantitation in cross-sectional and longitudinal analyses

**DOI:** 10.1101/2023.07.03.23292155

**Authors:** Gihan P. Ruwanpathirana, Robert C. Williams, Colin L. Masters, Christopher C. Rowe, Leigh A. Johnston, Catherine E. Davey

**Affiliations:** Department of Biomedical Engineering, The University of Melbourne, Melbourne, VIC, Australia; Melbourne Brain Centre Imaging Unit, The University of Melbourne, Melbourne, VIC, Australia; Florey Institute of Neurosciences and Mental Health, The University of Melbourne, Melbourne, VIC, Australia; The Australian Dementia Network (ADNET), Melbourne, Australia; Department of Molecular Imaging & Therapy, Melbourne, Austin Health, VIC, Australia

**Keywords:** Amyloid-β, reconstruction parameters, [^18^F] florbetapir, PSF, TOF, SUVR, CL

## Abstract

Amyloid beta (Aβ) accumulation in Alzheimer’s disease (AD) is typically measured using standardized uptake value ratio (SUVR) and the Centiloid scale (CL). The low spatial resolution of PET images is known to degrade quantitative metrics due to the partial volume effect (PVE). This paper examines the impact of spatial resolution, as determined by the reconstruction configuration, on the Aβ-PET quantitation in both cross-sectional and longitudinal data.

**Methods:** **Cross-sectional Study**-89 subjects with [^18^F]-florbetapir scans (44 Aβ-, 45 Aβ+) were reconstructed using 69 reconstruction configurations. For each reconstruction, Aβ SUVR was calculated and the spatial resolution was calculated as full-width-at-half-maximum (FWHM) using the barrel phantom method (Lodge *et al*, 2018). The change of SUVR and the effect size of the difference in SUVR between Aβ- and Aβ+ groups with FWHM were examined.

**Longitudinal study**-79 subjects (46 Aβ-, 33 Aβ+) with three [^18^F]-flutemetamol scans were analysed. All scans were reconstructed using low-, medium- and high-resolution reconstruction configurations and Aβ CLs were calculated. Since linear Aβ accumulation was assumed over a 10-year interval, for each reconstruction configuration, Aβ accumulation rate differences (ARD) between the second and first periods were calculated for all the subjects and compared. Zero ARD was used as a consistency metric. The number of Aβ-accumulators was also used to compare reconstruction configurations.

**Results:** **Cross-sectional-**SUVRs in both Aβ- and Aβ+ groups were impacted by the FWHM of the reconstruction method; Aβ- SUVRs increased for FWHM ≥ 4.5 mm, while Aβ+ SUVRs decreased across the FWHM range. High-resolution reconstructions provided the best statistical separation between groups.

**Longitudinal study**-In the Aβ-group, the median ARD of low-resolution reconstructed data was greater than zero whereas the ARDs of higher-resolution reconstructions were not significantly different to zero, indicating less consistent rates in the low-than the higher-resolution data. Higher-resolution reconstructions identified 10 additional Aβ-accumulators in the Aβ-group, resulting in a 22% increased group size compared to the low-resolution reconstructions. Higher-resolution reconstructions reduced the average CL values of the negative group by 12 points.

**Conclusions:** High-resolution PET reconstructions, inherently less impacted by PVE, may improve Aβ-PET quantitation in both cross-sectional and longitudinal data. In the cross-sectional analysis, separation of Aβ groups’ SUVRs increased with spatial resolution. Longitudinal analysis showed better Aβ accumulation consistency in higher-resolution compared to low-resolution reconstructions. The identification of more Aβ-accumulators from the higher-resolution reconstruction may be helpful in early-stage AD therapies.

## INTRODUCTION

Amyloid beta (Aβ) accumulation in the brain is a pathological indicator of Alzheimer’s disease (AD) that can be imaged using positron emission tomography (PET). Second-generation PET radiotracers, such as [^18^F]-florbetaben, [^18^F]-florbetapir and [^18^F]-flutemetamol, have been designed for on-target binding of Aβ plaques, enabling better diagnosis, management, and treatment of AD patients (*1,2*). The extent of Aβ-PET deposits is most commonly measured using scaled variants of the standardized uptake value (SUV), such as the standardized uptake value ratio (SUVR) and the Centiloid scale (CL) (*2,3*).

PET imaging has a low spatial resolution relative to other imaging modalities due to factors including the positron range of the radioisotope, photon scattering and hardware-specific limitations (*4*). The low resolution renders PET imaging particularly susceptible to the partial volume effect (PVE), in which quantitative PET metrics are degraded by the presence of multiple tissue types within a single voxel (*5*). Although the highest achievable spatial resolution of the PET image is determined by the scanner hardware, reconstruction algorithms and associated parameters are often chosen to provide lower resolution to maintain spatial noise variance at clinically accepted values (*6*). The ordered subset expectation maximization (OSEM) algorithm, characterised by the numbers of subsets and iterations, produces reconstructed images that are prone to increased spatial noise variance at high iteration numbers (*7*); this can be reduced by early termination of the iterative loop and post-reconstruction smoothing, at the cost of reduced spatial resolution and increased PVE (*6*–*8*).

The noise and spatial resolution of PET images has been improved by the use of time-of-flight (TOF) and point-spread-function (PSF) information in the reconstruction process (*9*). TOF incorporates the difference in detector arrival time between two photons into the reconstruction process, enabling both faster reconstruction convergence and reduced image noise (*10*). The PSF was introduced to improve the spatial resolution of PET images by incorporating the scanner response resolution into the reconstruction algorithm (*4,11,12*). In altering the quality of PET images, these reconstruction improvements impact the quantitative metrics derived from the resultant images (*11*–*13*).

Studies have examined the impact of the choice of the reconstruction algorithm and associated parameter configurations on tumour PET quantitation (*11*–*15*). However, only a few studies have examined the effect of reconstruction on neuroimaging PET quantitation. These studies were primarily based on phantom scans, often using a small subject cohort usually saved for retrospective reconstruction (*16*–*18*). A notable exception is a recent study examining the impact of reconstruction parameters on [^18^F]-FDG and [^18^F]-flutemetamol scans of AD subjects (*21*). This cross-sectional study showed a significantly lower Aβ SUVR in the AD control group with PSF-enabled reconstruction compared to OSEM reconstruction. In contrast, in the AD patient group, there was no significant difference in Aβ SUVR with PSF-enabled reconstruction. The study used a limited number of reconstructions, modifying the choice of the reconstruction algorithm and not the subset and iteration parameter choices. Studies examining the impact of reconstruction on longitudinal metrics of Aβ, and the varied impact of reconstruction on Aβ- and Aβ+ cohorts, are absent from the literature.

To examine the impact of PET reconstruction on longitudinal Aβ-PET measures in AD, it is necessary to establish an expectation of how Aβ-PET measures will change over time. Aβ accumulation is known to follow a sigmoidal pattern over decades of Aβ deposition (*20*). However, for longitudinal studies of relatively short duration, a linear Aβ accumulation rate can be assumed (*20*–*22*).; this assumption enables examination of the impact of reconstruction on Aβ quantitation over short periods of time.

In this study, we examined the impact of the spatial resolution of PET images, as determined by the choice of the reconstruction algorithm and associated parameters, on measures of AD progression, including Aβ SUVR and Aβ CL values. Both cross-sectional and longitudinal Aβ-PET datasets were reconstructed with a range of configurations and evaluated according to their impact on (i) cross-sectional Aβ SUVR values, and the subsequent separation of SUVR values between Aβ- and Aβ+ groups, and (ii) the estimated rate of Aβ accumulation over time.

## MATERIALS AND METHODS

### PET/CT scanner specifications

Subjects and phantoms were scanned on a Siemens Biograph mCT 128 PET scanner at the Melbourne Brain Centre Imaging Unit, the University of Melbourne. All data were recorded in three dimensions and saved in list-mode before reconstruction.

### Subject scanning

#### Cross-sectional study

A total of 89 subjects were selected from the Australian Imaging, Biomarker and Lifestyle study (AIBL) study. All subjects were scanned for Aβ by injecting [^18^F]-florbetapir radiotracer 50 minutes prior to 20 minutes of continuous scanning (Table 1).

**Table 1:** Comparison of resolution effects: MSE between CL and linearly predicted CL, and number of identified Aβ accumulators.

| Reconstruction type<br>(Algorithm,<br>resolution) | Mean<br>absolute error<br>(Aβ-) | Mean<br>absolute error<br>(Aβ+) ) | Number of<br>identified Aβ<br>accumulators<br>(Aβ-) | Number of<br>identified Aβ<br>accumulators<br>(Aβ+) ) |
| --- | --- | --- | --- | --- |
| Low resolution<br>(OP, 7.05 mm) | 2.23 ± 2.85 | 9.35 ± 16.68 | 17 | 26 |
| Medium resolution<br>(OPTOF, 4.55 mm) | 1.30 ± 1.65 | 9.39 ± 17.04 | 27 | 26 |
| High resolution<br>(PSFTOF, 3.05 mm) | 1.68 ± 2.19 | 13.19 ± 27.23 | 27 | 25 |

#### Longitudinal study

From the AIBL FLUTE longitudinal study, 79 subjects each with three longitudinal data points were selected. Subjects were injected with [^18^F]-flutemetamol radiotracer 50 minutes prior to 20 minutes of continuous scanning. The first and second follow-up scans were acquired 1.61 ± 0.14 and 3.21 ± 0.21 years after the baseline scan, respectively.

### Image Reconstruction

#### Spatial resolution calculation

A cylindrical phantom was used to measure the axial and radial PET spatial resolution of a given reconstruction configuration by calculating the full width at half maximum (FWHM) along each direction as proposed by Lodge *et al* (*8*). The phantom was filled with 64.7 MBq of ^18^F, and placed at the centre of the scanner FOV, with the long axis parallel to the axis of the scanner. One end of the phantom was lifted to create a small angle with the scanner axis, in order that the phantom edge intersects the image matrix obliquely in different slices. The tilted phantom was scanned for 60 minutes. Each phantom scan was reconstructed with a set of reconstruction configurations and uploaded to the Society of Nuclear Medicine and Molecular Imaging’s (SNMMI) phantom analysis toolkit (http://www.snmmi.org/PAT) to calculate axial and radial resolutions of each configuration in FWHMs (*8*). The average of the axial and radial resolution was the FWHM associated with the given reconstruction configuration.

#### Cross-sectional study

Each subject’s data were reconstructed using a set of reconstruction algorithms and parameter configurations chosen to cover a range of spatial resolutions. The FWHM associated with each reconstruction configuration was calculated using the cylindrical phantom method described above. Three reconstruction algorithms were used: Ordinary Poisson-OSEM (OP), OSEM+TOF (OPTOF) and OSEM+TOF+PSF (PSFTOF), each with two parameters: the number of OSEM iterations, *i*, and the number of subsets, *s*. The set of subsets and iterations differed between algorithms due to the parameter limitations of the scanner software. Consequently, OP, OPTOF and PSFTOF were implemented using each of six possible iterations, *i* ∈ {2,4,6,8,10,12}, with *s* = 21 for OPTOF and PSFTOF and *s* = 24 for OP. Additional OP reconstructions were performed using *i* ∈ {4,6,8,10,12} with *s* = 4. Post-reconstruction Gaussian filters of sizes 0 mm, 1 mm and 5 mm were applied. Hence, a given reconstruction configuration was comprised of the algorithm type, the number of subsets, the number of iterations, and the post-reconstruction smoothing size. In total, 69 reconstruction configurations were applied per dataset.

#### Longitudinal study

To evaluate the effect of the image spatial resolution on longitudinal Aβ-PET quantitation, each subject’s scan was reconstructed using three reconstruction choices representing three distinct resolution categories: low (FWHM=7.05 mm), medium (FWHM=4.55 mm) and high (FWHM=3.05 mm) resolution. The configuration details of each category were as follows: the 7.05 mm FWHM low-resolution reconstruction was implemented using OP (*i*=4, *s*=4, 2 mm post-reconstruction Gaussian smoothing); the 4.55 mm FWHM medium resolution using OPTOF (*i*=4, *s*=21, 0 mm post-reconstruction Gaussian smoothing); and the 3.05 mm FWHM high-resolution configuration implemented using PSFTOF (*i*= 4, *s*=21, 0 mm post-reconstruction Gaussian smoothing).

Both cross-sectional and longitudinal reconstructions were computed using the Siemens’ e7 toolbox with attenuation, decay, scatter and random corrections. Cross-sectional datasets were reconstructed at a voxel size of 4.07 mm × 4.07 mm × 2.01 mm and dimensions of 200 × 200 × 109 voxels. Longitudinal datasets were reconstructed at a voxel size of 1.99 mm × 1.99 mm × 3.00 mm and dimensions of 256 × 256 × 148 voxels.

### Data analysis

#### FWHM variation

To analyse the impact of reconstruction configurations on spatial resolution, phantom-derived FWHMs were used to evaluate the change of FWHM with reconstruction algorithm and iteration update (*i*×*s*) (**Supplementary Figure S1**).

#### Cross-sectional study

Each image reconstruction was uploaded to CapAIBL (http://milxcloud.csiro.au) to generate Aβ SUVR values, estimated by calculating tracer retention inside a neocortical mask of known Aβ accumulation brain regions (*23*). The Aβ positivity of the subjects was determined visually by an expert, separating subjects into Aβ- and Aβ+ groups. SUVR values were used in the cross-sectional study as CL values from CapAIBL were not available when the data was processed.

Hereinafter, this paper uses SUVR to denote the Aβ SUVR value generated by the CapAIBL software.

The effect of reconstruction on SUVR was examined by observing the change in mean SUVR values as a function of spatial resolution, quantified by FWHM, in both Aβ- and Aβ+ groups.

The impact of post-reconstruction smoothing on SUVR was analysed by calculating the mean pairwise SUVR difference between reconstructions with the same iteration and subset parameters but with differing post-reconstruction smoothing, of either 5 mm or 0 mm. This was evaluated separately across Aβ- and Aβ+ cohorts. The effect size of mean SUVR separation between the Aβ- and Aβ+ groups was calculated using Cohen’s d, as a function of the FWHM associated with reconstruction configurations.

To visualize the impact of spatial resolution on the separation between Aβ - and Aβ+ groups, the reconstruction configuration with the largest effect size (group separation) was compared with a typical clinical reconstruction configuration of OP with 4 iterations, 4 subsets, and 5 mm post-reconstruction Gaussian smoothing. For each reconstruction configuration, SUVR values of both Aβ- and Aβ+ groups were tested for Gaussianity, and subsequently fitted with Gaussian distributions, enabling a comparison of SUVR group separation between the two reconstruction configurations, via t-tests on estimated means. The SUVR means of high-resolution reconstructions in Aβ- and Aβ+ groups were compared with the relevant SUVR mean of clinical reconstruction configuration with right-tail and left-tail t-tests, respectively. A left-tailed F-test was used to compare the SUVR variances of high-resolution reconstructions in both Aβ- and Aβ- groups with the respective SUVR variance of the clinical reconstruction configuration.

#### Longitudinal study

To ascertain the effect of PET reconstruction on longitudinal Aβ quantitation, CL values were used to measure global Aβ-PET uptake, estimated by uploading each reconstructed image volume to CapAIBL. Subjects were allocated into Aβ– and Aβ+ groups, where the Aβ positivity of a subject was determined using the CL estimate of the low-resolution OP reconstructed baseline scan, with a threshold CL ≥ 20 (Table 1).

Based on known properties of the Aβ accumulation process, over a 10-year interval a linear Aβ accumulation rate can be assumed, resulting in a consistent accumulation across the 10 year period (*21*). Since there were three scans per subject within 5 years, well within the 10-year duration of linear Aβ accumulation, Aβ accumulation rates in both the first interval and second inter-scan intervals were computed and tested for conformity with the linearity assumption. Indeed, the linearity of the Aβ accumulation process was used to derive two surrogate metrics to analyse the impact of reconstruction configurations on longitudinal Aβ CL quantitation.

The first surrogate metric is the Aβ rate difference (ARD) between the second and first intervals for each subject and each reconstruction configuration. Given a linear Aβ accumulation assumption over the longitudinal study period, ARD = 0 indicated a consistent accumulation rate over the three-time points. The Wilcoxon signed-rank test was used to test whether the median ARD of each reconstruction was statistically significantly different from zero in both Aβ- and Aβ+ groups; for low- and high-resolution reconstruction configurations, right-tail Wilcoxon signed-rank tests were used in both Aβ groups, where, for the medium resolution configuration, left- and right-tail Wilcoxon signed-rank tests were used in Aβ- and Aβ+ groups, respectively. We decided on the use of right- or left-tail Wilcoxon signed-rank tests depending on the sign of the median ARD value. A right-tail Mann-Whitney *U* test was also carried out on both Aβ- and Aβ+ groups to test whether there was a difference in median ARD across reconstruction configurations.

For each subject and for each of the three reconstruction choices, a linear model was fitted across the three-time points. The average mean squared error (MSE) of each model was used as a second surrogate marker of longitudinal consistency of Aβ accumulation. Wilcoxon signed-rank tests were carried out to test whether the median accumulation slopes of the fitted linear models were statistically significantly different from zero as it was an indication of the number of generated positive slopes; right-tail Wilcoxon signed-rank tests were used across all the reconstructions in both Aβ groups, except for low-resolution reconstruction configuration in the Aβ- group, which used left-tail Wilcoxon signed-rank test. We decided on the use of right- or left-tail Wilcoxon signed-rank tests depending on the sign of the median accumulation slope. The median accumulation slopes of the three reconstruction configurations were also compared using the left-tail Mann-Whitney *U* test. All tests were performed on Aβ- and Aβ+ groups separately.

In addition to the above statistical tests, the number of Aβ accumulators, identified as subjects with positive accumulation slopes, was calculated as a third comparison measure of reconstruction configurations.

## RESULTS

Demographics of subjects used in the cross-sectional and longitudinal studies are given in Table 1. The cross-sectional study had a balanced dataset with similar female percentages and ages in both Aβ- and Aβ+ groups. The longitudinal study had more Aβ-subjects whilst the Aβ+ group was older than the Aβ-group.

### Cross-sectional study

SUVRs in both Aβ- and Aβ+ groups were impacted by the spatial resolution of the reconstruction configuration (Figure 1). The standard errors of SUVR in the Aβ- group were small compared to the Aβ+ group for all reconstructions (Figure 1A, B). In the Aβ-group, no notable SUVR differences were seen across low FWHM reconstructions with 0 mm and 1 mm smoothing; SUVR values started to increase at an FWHM value of approximately 4.5 mm (Figure 1A). Reconstructions with 5 mm smoothing showed the same trend, but with a plateaued region from 5.2 mm to 7 mm FWHM, followed by an increase in mean SUVRs. Application of post-reconstruction smoothing shifted the no-smoothing reconstruction data to the right without a notable SUVR difference Figure 1A red, blue and orange markers). In the Aβ+ group, there was no plateaued SUVR region as in the Aβ-group, with SUVR values decreasing across the full range of FWHM values (Figure 1B).

**Figure 1:**
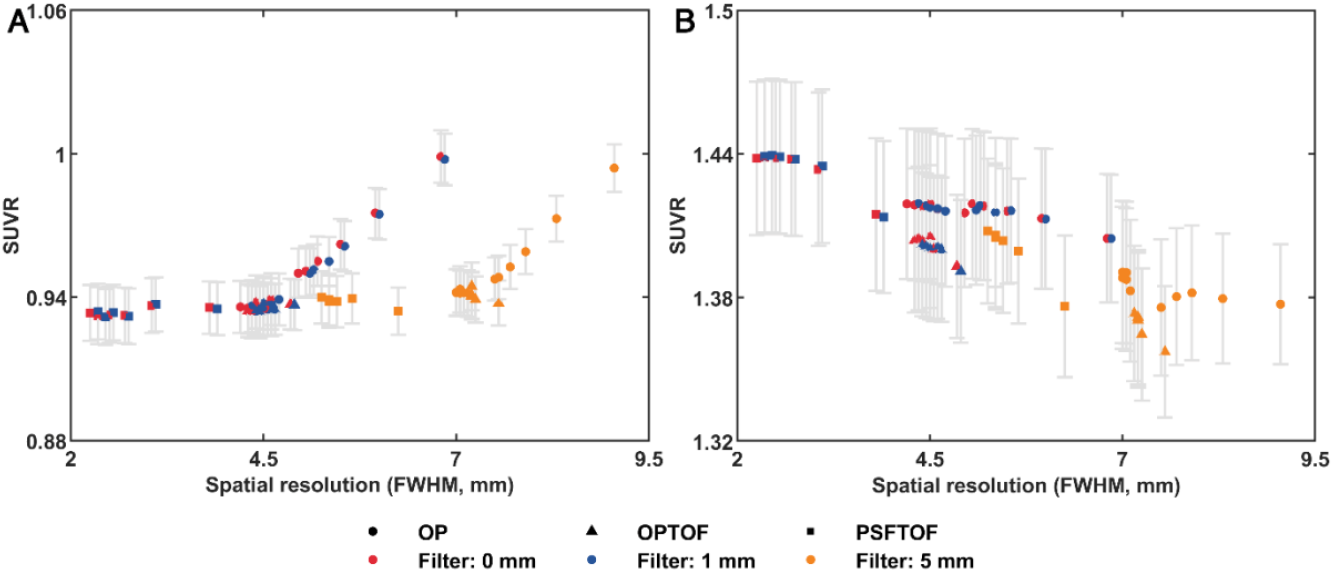
Spatial resolution versus Aβ SUVR. The impact of spatial resolution (FWHM), as determi11ed by reconstruction configurations, on mean Aβ SUVR in A) Aβ- and **B)** Aβ+ groups. Each data point represents the mean Aβ SUVR across all subjects in a group reconstructed using the same algorithm, number of iterations, subsets and post-reconstruction Gaussian smoothing. The spatial resolution of a data point vvas the calculated FWHM of a cylindrical phanto111 reconstructed using the same parameter settings (8). Reconstruction algorithms are designated by marker shape; OP (circles), OPTOF (triangles), and PSFTOF (squares). Tl1e filter value in the legend denotes the FWHM of thepost-reconstruction Gaussian smoothing. Grey whiskers = standard error.

Figure 1 shows mean SUVR values with varying post-reconstruction smoothing filters applied to each reconstruction configuration. For the Aβ-cohort, mean SUVR appears to remain constant, despite the consequent shift in FWHM due to post-reconstruction smoothing, with each data set showing the same range and trend (Figure 1A). However, Figure 1B indicates that post-reconstruction smoothing may impact mean SUVR for the Aβ+ cohort, in the noticeable shift between the red/blue and orange markers. This issue was examined in more detail in the supplementary material; Supplementary Figure S2 shows changes in mean Aβ SUVR for each reconstruction group after the images are smoothed with a 5 mm post-reconstruction Gaussian filter. As per Supplementary Figure S2, post-reconstruction smoothing impacted the SUVR of the Aβ+ group, with the application of a 5 mm smoothing filter reducing the Aβ+ group’s mean SUVR value for every reconstruction configuration (Supplementary Figure S2, red markers). However, for the Aβ-group, the change in mean SUVR after the application of a 5 mm smoothing filter was negligible for all reconstruction configurations (Supplementary Figure S2, blue markers).

To analyse the impact of the reconstruction configuration on the separation between the SUVRs of the Aβ-and Aβ+ groups, the effect size of the difference in mean SUVR between the groups as measured by Cohen’s d was compared against the FWHM value of the reconstruction method (Figure 2). The maximum effect size between the groups was achieved by PSFTOF (10*i*, 21*s*) with 1 mm smoothing, with similar effect sizes for all PSFTOF reconstructions with 0 mm and 1 mm smoothing. Effect sizes decreased with FWHM value, irrespective of the reconstruction method and the level of smoothing used.

**Figure 2:**
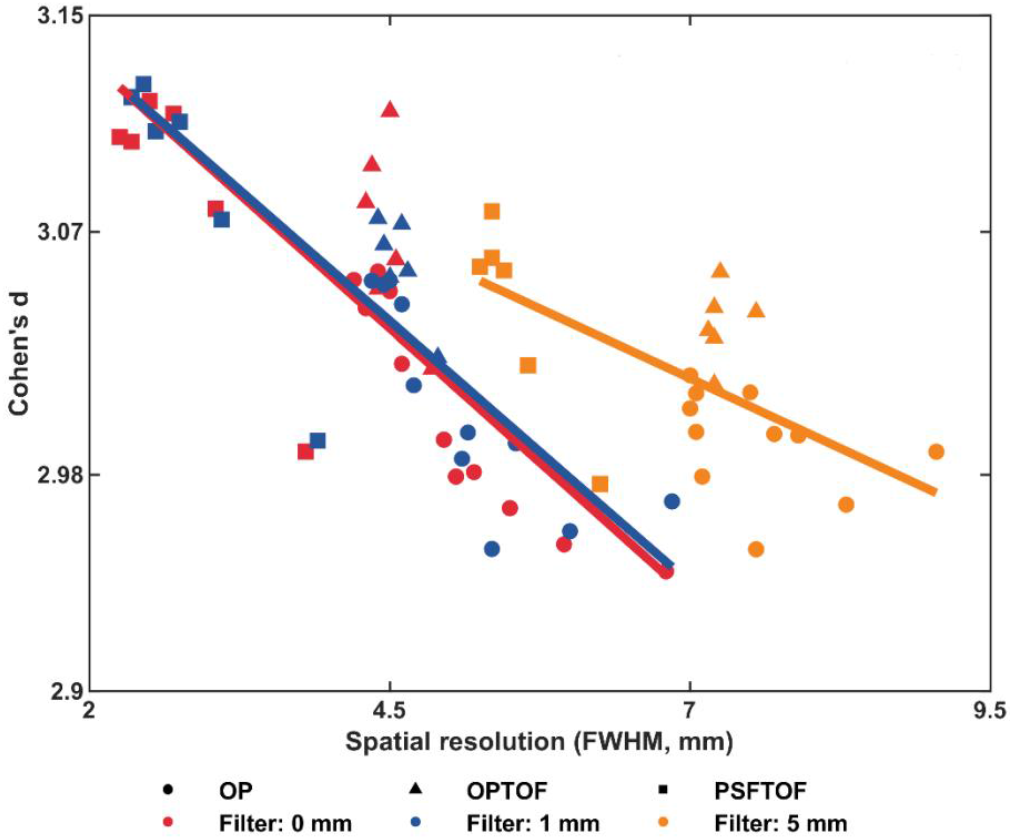
Spatial resolution versus the effect size of the difference in mean SUVR between Aβ- and Aβ+ groups as measured by Cohen’s d. The impact of the spatial rcsolution of each reconstuction configuration on the effect size of the difference in mean SUVR between Aβ+ and AP groups. Correlations between FWHM and Cohen’s d for different post reconstruction Gaussian smoothing amounts are -0.818, -0.862, and -0.622, for 0 mm, 1 mm and 5 mm filters, respectively. Lines of best fit are depicted by red (0 mm), blt1e (1 mm) and orange (5 mm).

The separation between Aβ- and Aβ+ groups was dependent on the resolution of the reconstruction configuration. The difference in separation between Aβ+ and Aβ-groups with reconstruction configuration is shown in Figure 3 by using a standard clinical reconstruction and a high-resolution reconstruction, resulting in the largest separation between the cohorts. Although both the standard clinical reconstruction and the high-resolution reconstruction separated the Aβ+ and Aβ-groups significantly (*p*<0.00005), the latter increased the dynamic range of SUVR values in both groups Figure 3C); the mean SUVR of the high-resolution reconstruction was significantly smaller (*p*=0.00006) than the standard reconstruction in Aβ- group where the mean SUVR of Aβ+ group trended towards a significantly larger value (*p*=0.07) in the high-resolution reconstruction compared to the standard reconstruction. Moreover, SUVR variances of the high-resolution reconstruction trended towards significantly larger values than the standard reconstruction in both Aβ-(*p*=0.15) and Aβ+ (*p*=0.05) groups.

**Figure 3:**
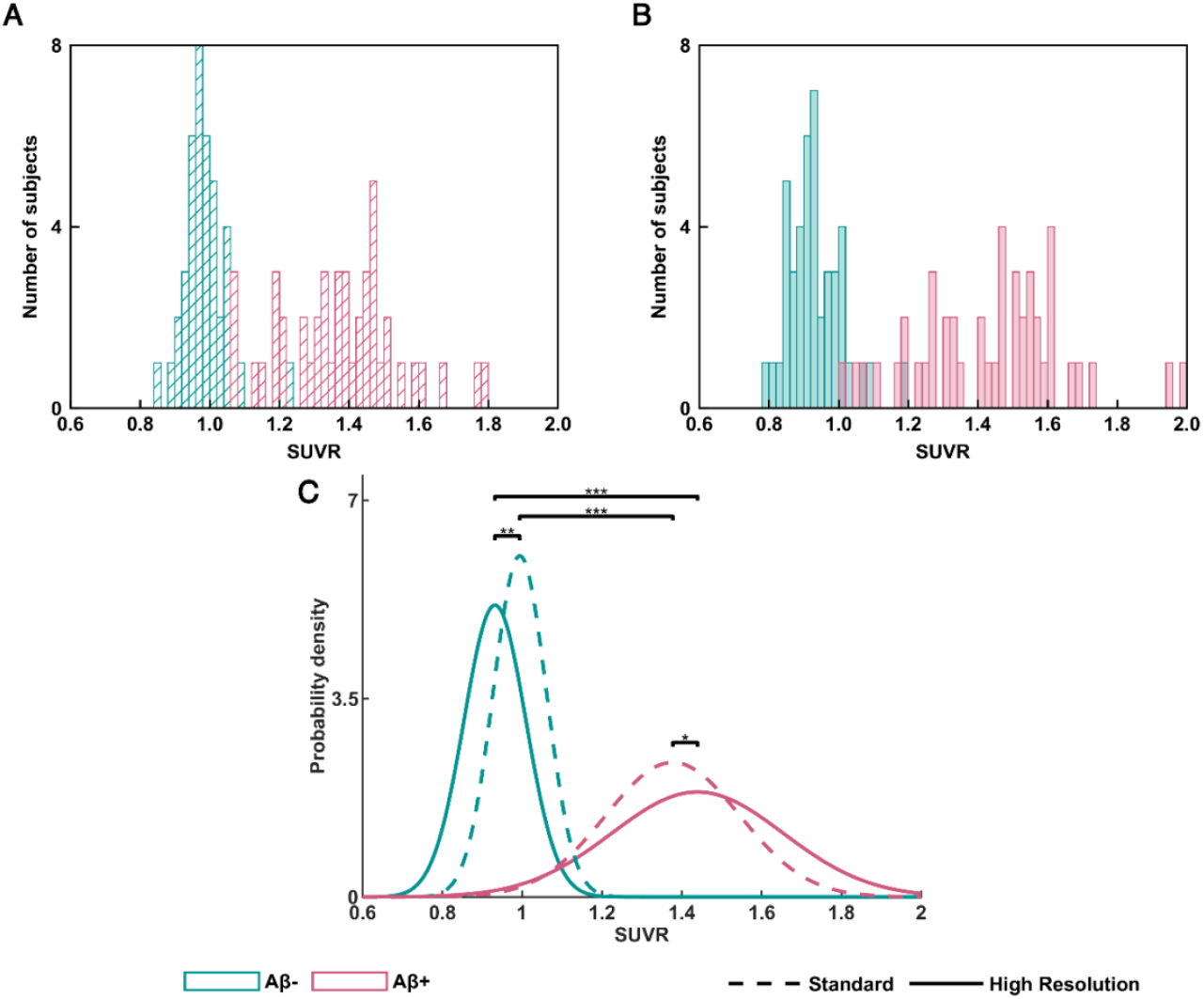
Separation between Aβ- and Aβ+ groups for different reconstructions. The separation between Aβ and Aβ+ groups using **A)** a clinical reconstruction configuration OP (4*i*, 4*s*) with 5mm smoothing and **B)** a high resolution reconstruction co11figuration, PSFTOF OP (10*i*, 21*s*) with 1mm smoothing, resulted in the largest separation between the Aβ- and Aβ+ cohorts. C) Fitted Gaussian distributions on S UYRs of both reconstruction configurations are overlaid for ease of visualisation. Annotations: * denotes a trend towards a significant difference in the mean (*p*=0.07) and variance (*p*=0.05), ** denotes a significant difference in the mean (*p*=0.00006) and a trend towards a significant difference in variance (*p*=0.15), while *** represents a significant difference between group means (*p*<0.00005).

### Longitudinal study

The consistency of longitudinal measurements of Aβ accumulation was analysed across reconstruction configurations by analysing the Aβ accumulation rate differences (ARDs) between the two periods, where ARD = 0 denotes the best level of consistency (Figure 4A, B). The data underlying these results is summarised in Supplementary Figures S3 and S4. In the Aβ-group, the median ARD of low-resolution reconstructed data trended towards a statistically significant value greater than zero (*p*=0.1) and the median ARD of both medium-and high-resolution reconstructions were not significantly different from zero. Higher-resolution reconstructions reduced the average CL values of the negative group by 12 points. In contrast, in the Aβ+ group, median ARD of both medium-resolution (*p*=0.006) and high-resolution (*p*=0.004) reconstructions were significantly greater than zero and the median ARD of low-resolution reconstructions trended towards a value significantly greater than zero (*p*=0.08). Consequently, medium- and high-resolution reconstructions resulted in more consistent Aβ-PET longitudinal data than the low-resolution reconstructions, especially in the Aβ-group.

**Figure 4:**
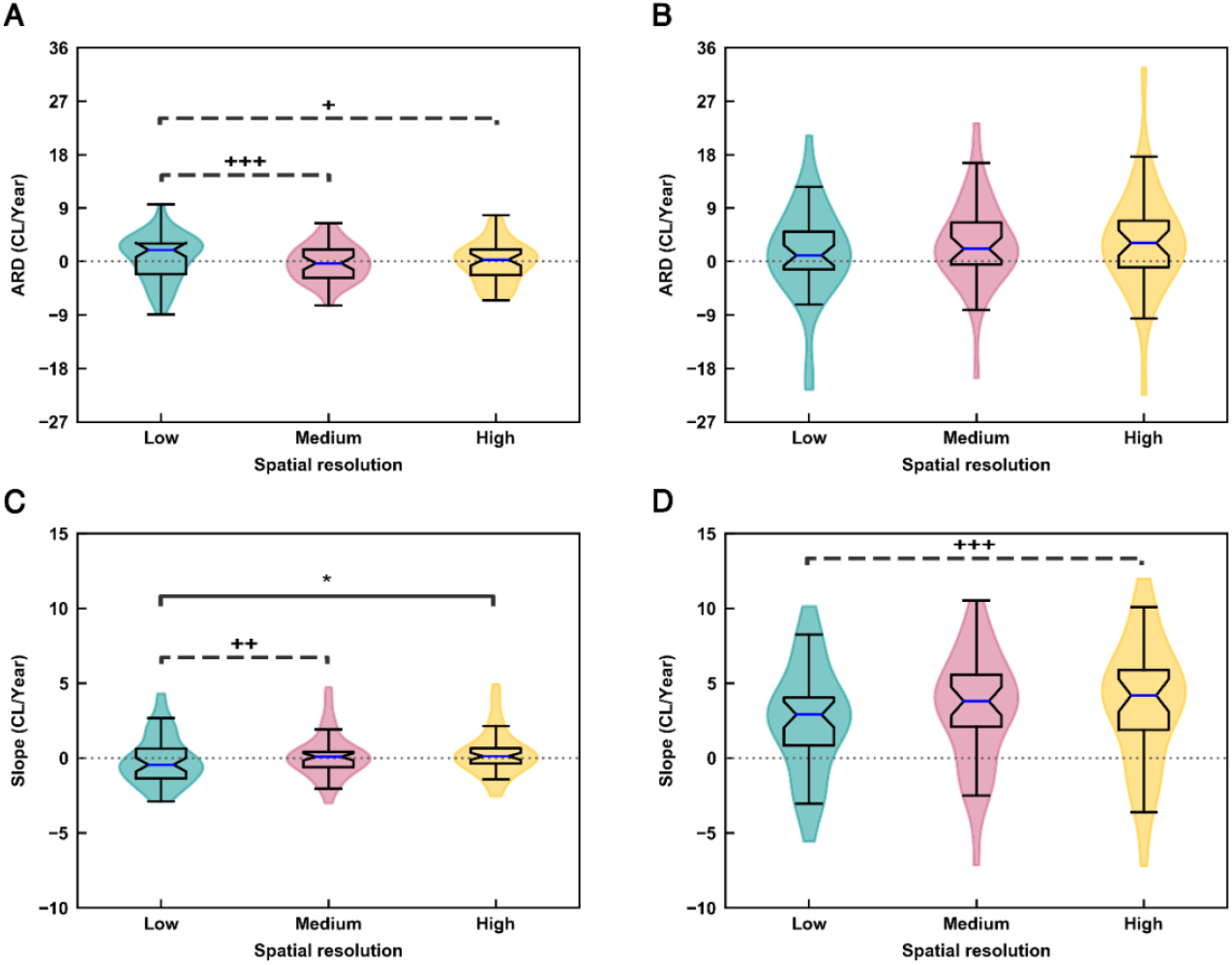
Comparison of accumulation rate differences (ARD) and fitted accumulation slopes across reconstructions. The ARDs between second and first-time intervals for three reconstructions: low-resolution OP (4*i*, 4*s*, 2 mm smoothing, FWHM 7.05mm), medium-resolution OPTOF (4*i*, 21*s*, Omm smoothing, FWHM 4.55 mm) and high-resolution PSFTOF (4*i*, 21*s*, 0mm s111oothing, FWHM 3.05mm) in A) Aβ- and **B)** Aβ+ groups. Aβ acctmulation slopes of longitudinal linear models for **C)** Aβ-and **D)** Aβ+ groups. In group ditlerence annotatios, the solid line indicates a signitficant median difference, the dotted line indicates a trend towards a significant median difference, * denotes *p*=0.03, + denotes *p*=0.08, ++ denotes *p*=0.07 and.+++ denotes *p*=0.06. The high and medium resolution reconstructions of the Aβ+ group show an average increase in CL/Year of 1.05 and 0.70 over the low-resolution reconstruction group, respectively.

The median ARD values were dependent on the resolution of the reconstruction configuration. In the Aβ-group, the median ARD of low-resolution reconstructions trended towards a significantly larger value than the ARD medians of medium-(*p*=0.06) and high-resolution (*p*=0.08) data (Figure 4A). However, significant changes in ARDs were not observed across the three reconstructions in the Aβ+ group (Figure 4B).

Slopes of linear models fitted to the longitudinal data were impacted by the spatial resolution of the reconstruction (Figure 4C, D). In the Aβ-group, the median slope of the low-resolution reconstructions was negative and trended toward a significantly non-zero value (*p*=0.1), whereas the median slope of the high-resolution reconstructions was positive and trended toward a significantly non-zero value (*p*=0.1). The positive median slope of the medium-resolution data was not significantly different from zero (*p*=0.4). In contrast, in the Aβ+ group, all median slopes of low-resolution (*p*<0.05), medium-resolution (*p*<0.05) and high-resolution (*p*<0.05) data were significantly larger than zero; the median slopes of the medium- and high-resolution reconstructions were greater than that of the low-resolution reconstructions. However, only the median slope of high-resolution reconstructions trended towards a significantly larger value (*p*=0.08) than the median slope of low-resolution reconstructions. Thus, high- and medium-resolution reconstructions produced more positive slopes than the low-resolution reconstruction.

We turn now to an analysis of the relative median accumulation slopes between low-, medium- and high-resolution reconstructions in both Aβ- and Aβ+ groups. In the Aβ- group, the median accumulation slope of high-resolution reconstruction data was significantly larger than the low-resolution case (*p*=0.03) and the median accumulation slope of medium-resolution trended towards a larger value (*p*=0.07) than the low-resolution case (Figure 4C). Notably, the difference in median accumulation slopes was not seen in the Aβ+ group, except for a trend toward a significant difference between high- and low-resolution reconstructions (*p*=0.06) (Figure 4D).

The MSE difference between the actual CL value and linearly predicted CL value after fitting a linear model to subjects’ longitudinal CL data was used as a proxy for the longitudinal consistency in Aβ accumulation. In the Aβ- group, medium-resolution and high-resolution reconstructions had smaller MSE values compared to low-resolution reconstructions (Table 1). However, in the Aβ+ group, mean errors were similar across all the reconstructions. Moreover, medium- and high-resolution reconstructions identified 10 more additional Aβ accumulators in the Aβ- group compared to low-resolution reconstruction data; percentage-wise, higher-resolution reconstructions showed a 22% increase in identifying Aβ accumulators. There was no notable difference in the number of accumulators in the Aβ+ group (Table 1). In summary, medium- and high-resolution data showed better longitudinal consistency in measured Aβ accumulation and identified more Aβ accumulators in the Aβ- group.

## DISCUSSION

This study has examined the impact of spatial resolution of PET images, as determined by the reconstruction configuration, on Aβ-PET quantitation. The choice of PET reconstruction algorithm, each with different parameter configurations, is known to influence the spatial resolution of the resulting images, and affect quantitative values of PET metrics due to partial volume effects (*16,24*). The impact of reconstruction parameter choices on quantitation has been extensively studied in the PET oncology domain (*11*–*15*). Few studies have evaluated the impact of PET reconstruction on Aβ-PET imaging. In the current study, we have examined the impact of spatial resolution, as determined by reconstruction parameters, on quantitation in both cross-sectional and longitudinal Aβ-PET imaging datasets.

The spatial resolution of the reconstruction methods, as quantified by phantom-derived FWHM values, was observed to impact SUVR quantitation of cross-sectional data in both Aβ- and Aβ+ groups. The Aβ+ group had larger standard errors due to a wider range of SUVR values compared to Aβ- group, as expected. An increase and a decrease in SUVR values were observed with the FWHM values of the reconstruction configurations in the Aβ- and Aβ+ groups, respectively. This may be due to the partial volume effect in white matter and grey matter of the brain; although current generation Aβ-PET radiotracers are designed to bind to grey matter structures, some non-specific binding occurs, especially in white matter structures (*2*). As stated in previous studies, Aβ-subjects show higher tracer retention in white matter structures than grey matter structures devoid of Aβ, whilst grey matter structures of Aβ+ subjects show higher or equal tracer retentions than white matter structures (*25*). In Aβ-subjects, partial voluming may have resulted in the high activity of white matter contaminating grey matter values, resulting in artificially increased SUVR values. The reverse effect from partial voluming is likely to have occurred in the Aβ+ group, with a dilution of grey matter SUVRs from adjacent lower activity tissue. As the level of PVE changes with the spatial resolution of the reconstruction configuration, so too does SUVR quantitation.

Our cross-sectional analysis demonstrated that although the application of smoothing changed the FWHM of the reconstruction, it did not have a significant effect on the SUVR values in the Aβ-group. In contrast, there was a marked SUVR reduction in the Aβ+ group after the application of post-reconstruction Gaussian smoothing. We conclude that post-reconstruction smoothing may hinder SUVR quantitation by yielding differential effects on Aβ- and Aβ+ groups. Past studies have also suggested that smoothing may reduce or remove small differences due to pathology (*26*). Therefore, it is worth reconsidering scanner harmonization methods that use post-reconstruction smoothing to match the spatial resolution as a mitigation of multicentre PET quantitation differences (*27*–*29*). A possible alternative for harmonizing scanners is to match the non-smoothed FWHM values between scanners by adjusting reconstruction parameters, such as the number of iterations and subsets. Further studies are needed to examine the potential of non-smoothed FWHM matching for multi-scanner harmonization.

High-resolution reconstructions resulted in a larger separation between Aβ- and Aβ+ groups while increasing the dynamic range of SUVR values; cross-sectional analysis showed that differences between Aβ- and Aβ+ groups decreased with increasing FWHM. This was a consequence of the opposing SUVR trend in the two Aβ groups, where SUVRs increased in the Aβ-group and decreased in the Aβ+ group with FWHM. This is in contrast to the recent study that reported similar Cohen’s-d values irrespective of the reconstruction method (*19*), however it is notable that a smaller cohort and a more limited range of reconstruction configurations were used, along with a different Aβ-PET radiotracer. Moreover, our cross-sectional results showed that reconstruction configurations with better resolution gave the minimum and maximum SUVR values in Aβ- and Aβ+ groups, respectively; this may be due to the inherently lower PVE associated with high-resolution reconstructions. These results suggest that we should re-examine the use of modern reconstruction configurations with better spatial resolution as a standard reconstruction protocol in Aβ-PET neuroimaging.

To the best of our knowledge, this is the first study examining the impact of spatial resolution on longitudinal Aβ-PET data. As shown by the ARD metric, we found that medium- and high-resolution reconstructions resulted in more consistent Aβ accumulations across the longitudinal time course than low-resolution reconstructions, particularly in the Aβ-group. This is a clinically important result, as longitudinal Aβ-PET data should match the assumed linearity property of the Aβ accumulation process over a short time interval. Moreover, it is of crucial importance to clinical management of early-stage AD therapies to identify Aβ accumulators at low Aβ levels, as AD prevention therapies require testing at the early AD stages. We have demonstrated that a 22% higher number of Aβ accumulators in the Aβ- group were found using medium- and high-resolution reconstruction configurations than the more typical clinical low-resolution reconstruction configuration.

In the longitudinal study, the subjects were binarized to Aβ- and Aβ+ groups using a clinically typical CL cut-off point of 20 (*21*). However, it is important to note that CL value are dependent on the reconstruction configuration employed, leading to variation in the cut-off point between Aβ- and Aβ+ groups. Future studies should re-examine the concept of a universal cut-off point and consider the alternative of using tailored cut-off points depending on the reconstruction configuration employed.

The limitations of the current study include: (i) Noise measurements were not analysed in the study as we were only concerned with global Aβ-PET quantitation that was calculated using a mean uptake in a large region of interest in the brain; (ii) A lower radioactivity dose than used in the clinical configuration was injected into the barrel phantom; (iii) CL values were not used in the cross-sectional analysis; (iv) Only two Aβ-PET radiotracers were used. The results may depend on the radiotracer as the non-specific binding may change across tracers. Irrespective of these limitations, we found that high-resolution, converged reconstructions were better for Aβ-PET quantitation than low-resolution reconstructions, in both cross-sectional and longitudinal studies. Newer generation PET/CT scanners with even higher resolution and higher sensitivity digital detectors are now available that are expected to increase these benefits even further. Future studies can be conducted to examine the feasibility of harmonization between scanners by matching the barrel phantom-derived spatial resolutions of reconstruction methods.

## CONCLUSION

High-resolution reconstructions, with inherently less partial volume effects, can improve both cross-sectional and longitudinal Aβ-PET data quantitation as demonstrated by the increased separation between Aβ- and Aβ+ groups in the cross-sectional analysis and more consistent Aβ accumulation in longitudinal data than low-resolution scans, in conjunction with the identification of more Aβ accumulators in the Aβ-group; these Aβ-PET quantitation improvements gained from high-resolution reconstructions are an important aspect of understanding AD progression and the management of early-stage AD therapies using PET imaging. Our results demonstrate that post-reconstruction smoothing may hinder SUVR quantitation, suggesting that the use of post-reconstruction smoothing as a scanner harmonization method may not fully achieve its intended outcome.

## Supporting information

Supplementary methods and results

## Data Availability

All data produced in the present study are available upon reasonable request to the Australian Imaging and Biomarkers Lifestyle Study.

## DISCLOSURE

The research was supported by NHMRC and NIH grants. No potential conflicts of interest relevant to this article exist.

The human scans were approved by the Austin health Human Research Ethics Committee (HREC/18/Austin/201) and all experiments were performed in accordance with relevant guidelines and regulations.

## ACKNOWLEDGMENTS

The authors acknowledge the facilities and scientific and technical assistance of the National Imaging Facility, a National Collaborative Research Infrastructure Strategy (NCRIS) capability, at the Melbourne Brain Centre Imaging Unit, University of Melbourne. The first author would also like to acknowledge the Rowden White scholarship for its assistance in his research.

## KEY POINTS

**Question:** Does the PET spatial resolution, as determined by the choice of reconstruction configuration affect the quantitation of cross-sectional and longitudinal Aβ-PET?

**Pertinent Findings:** In the cross-sectional analysis, high-resolution reconstructions gave the lowest and highest Aβ SUVRs in Aβ- and Aβ+ groups, respectively, while improving the separation between the groups. High-resolution reconstructions improved the longitudinal consistency of measured Aβ accumulation and identified more Aβ accumulators.

**Implications for patient care:** High-resolution scans can improve both cross-sectional and longitudinal Aβ-PET data quantitation, which will assist in understanding AD progression and management of AD therapies in the early stages.

