## Supplementary methods and results for "Impact of PET reconstruction on Aβ-amyloid quantitation in cross-sectional and longitudinal analyses"

**SUPPLEMENTARY MATERIALS**

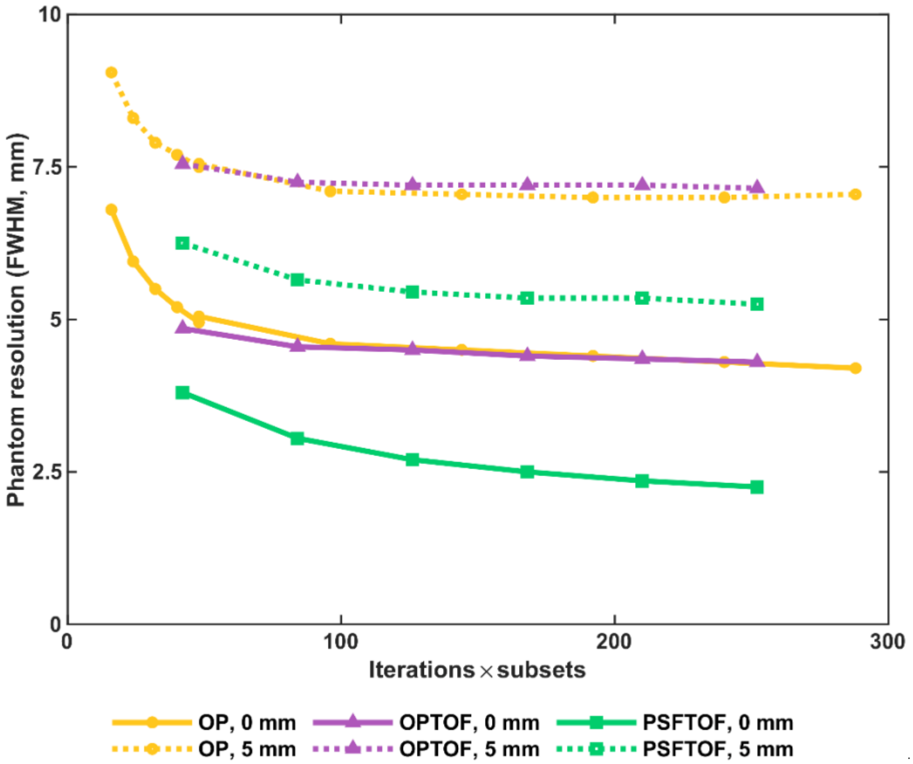


**FIGURE S1: Iteration update vs phantom resolution**. Comparison of the barrel phantom calculated

spatial resolution (FWHM) with the iteration update while using a post-reconstruction Gaussian smoothing of 0 mm and 5 mm.

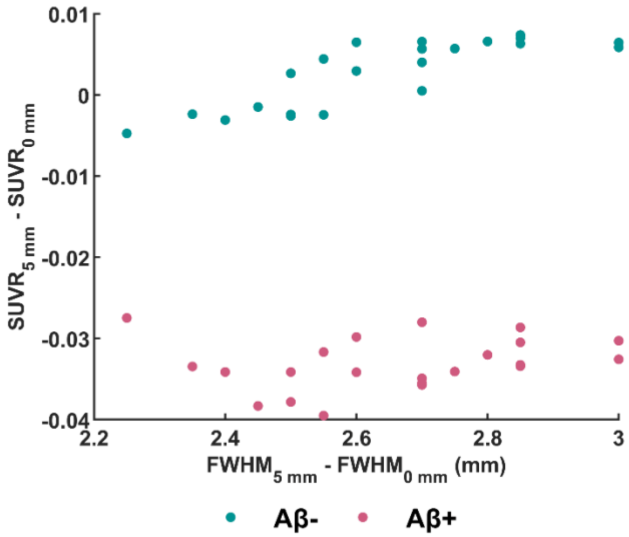


**FIGURE S2: The impact of post-reconstruction smoothing on Aβ SUVR.** Change in mean SUVR for each reconstruction group after the application of a 5 mm post-reconstruction Gaussian smoothing. Aβ- and Aβ+ groups are shown separately. Note: Error bars are not included as they are too small to be legible.

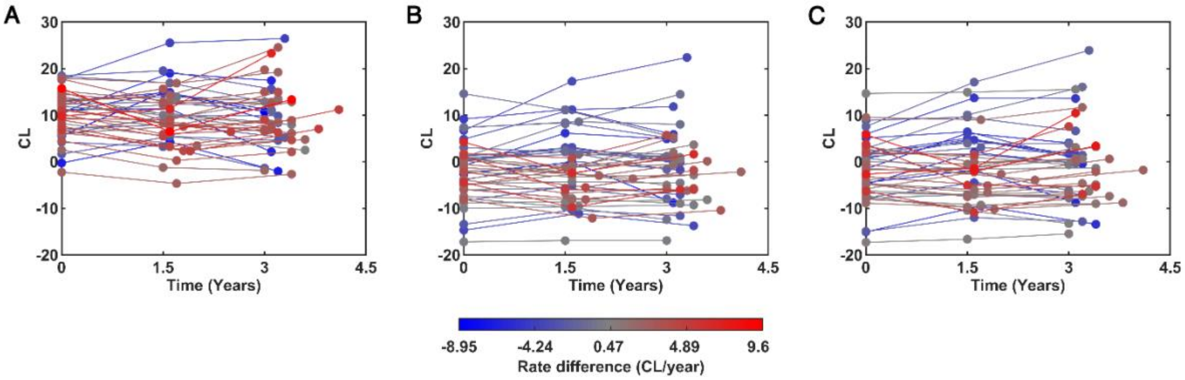


**FIGURE S3: Summary of Aβ- longitudinal data reconstructed with low-, medium- and high-resolution reconstruction configurations**. The impact of PET spatial resolution on the Aβ accumulation rate difference

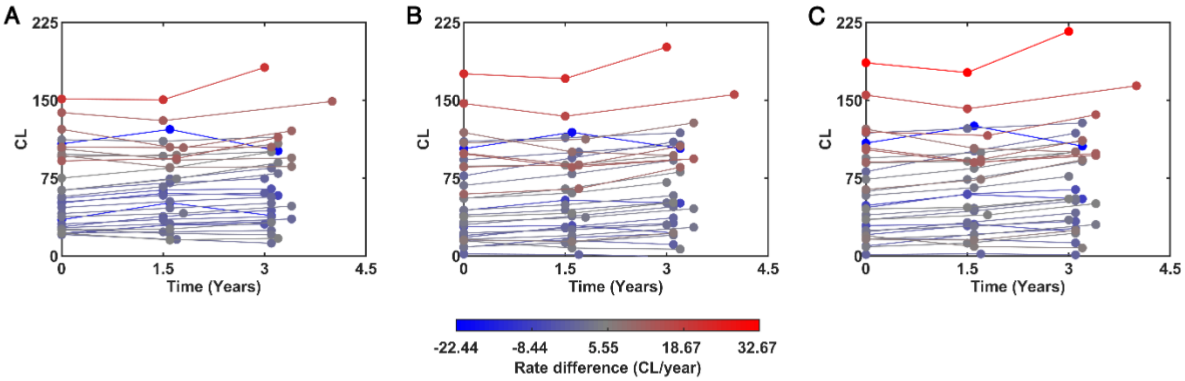


**FIGURE S4: Summary of Aβ+ longitudinal data reconstructed with low-, medium- and high-resolution reconstruction configurations**. The impact of PET spatial resolution on the Aβ accumulation rate difference.

**TABLE S1:** Demographics of Cross-sectional and Longitudinal study datasets.

| Study Type | Cross-sectional | | Longitudinal | |
|---|---|---|---|---|
| **Aβ group** | Aβ- | Aβ+ | Aβ- | Aβ+ |
| **Sample size** | 44 | 45 | 46 | 33 |
| **Sex. F (%)** | 26 (59) | 26 (58) | 22 (47.83) | 13 (39.39) |
| **Age ± SD** | 71 ± 3.77 | 70.78 ± 3.96 | 71.91 ± 4.51 | 74.00 ± 5.59 |
| **% Right hand** | 91 | 91 | 89* | 84* |
| **% APOE4** | 21* | 70 ** | 15 | 39 |
| **CU, MCI, AD** | 41, 0, 0*** | 37, 4, 0**** | 41, 4, 1 | 24, 4, 4* |
| **MMSE ± SD** | 27.9 ± 1.22 | 27.71 ± 1.33 | 29.09 ± 0.98 | 27.52 ± 2.83 |
| **CDR ± SD** | 0 ± 0* | 0.03 ± 0.13 | 0 .05 ± 0.19 | 0.17 ± 0.30 |
| **CDR-SoB ± SD** | 0.01 ± 0.08* | 0.09 ± 0.22 | 0.18 ± 1.03 | 0.79 ± 1.90 |

Demographics marked with * are missing values from some subjects, where the number of * corresponds to the number of subjects missing. F = Female, SD = Standard Deviation, APOE4 = Apolipoprotein E4, CU = Cognitively unimpaired, MCI = Mild Cognitively Unimpaired, AD = Alzheimer's Disease, MMSE = Mini-Mental State Examination, CDR = Clinical Dementia Rating, CDR-SoB = Clinical Dementia Rating Scale Sum of Boxes.

**TABLE S2:** Reconstruction configurations used in the cross-sectional analysis and their relevant Aβ-PET SUVR and FWHM values.

| Recons-truction method | Iterations | Subsets | Filter size (mm) | FWHM (mm) | Mean SUVR of Aβ- group | SD of Aβ- group | Mean SUVR of Aβ+ group | SD of Aβ- group |
|---|---|---|---|---|---|---|---|---|
| PSFTOF | 12 | 21 | 0 | 2.25 | 0.933 | 0.077 | 1.438 | 0.215 |
| PSFTOF | 10 | 21 | 0 | 2.35 | 0.932 | 0.079 | 1.439 | 0.216 |
| PSFTOF | 12 | 21 | 1 | 2.35 | 0.934 | 0.077 | 1.439 | 0.214 |
| PSFTOF | 10 | 21 | 1 | 2.45 | 0.932 | 0.077 | 1.439 | 0.215 |
| PSFTOF | 8 | 21 | 0 | 2.5 | 0.933 | 0.077 | 1.438 | 0.215 |
| PSFTOF | 8 | 21 | 1 | 2.55 | 0.934 | 0.077 | 1.439 | 0.216 |
| PSFTOF | 6 | 21 | 0 | 2.7 | 0.932 | 0.077 | 1.438 | 0.215 |
| PSFTOF | 6 | 21 | 1 | 2.75 | 0.932 | 0.077 | 1.438 | 0.216 |
| PSFTOF | 4 | 21 | 0 | 3.05 | 0.937 | 0.076 | 1.434 | 0.214 |
| PSFTOF | 4 | 21 | 1 | 3.1 | 0.937 | 0.075 | 1.435 | 0.215 |
| PSFTOF | 2 | 21 | 0 | 3.8 | 0.936 | 0.073 | 1.415 | 0.214 |
| PSFTOF | 2 | 21 | 1 | 3.9 | 0.935 | 0.073 | 1.414 | 0.213 |
| OP | 2 | 24 | 0 | 4.2 | 0.936 | 0.073 | 1.419 | 0.211 |
| OPTOF | 12 | 21 | 0 | 4.3 | 0.934 | 0.073 | 1.404 | 0.202 |
| OP | 10 | 24 | 0 | 4.3 | 0.936 | 0.072 | 1.419 | 0.212 |
| OPTOF | 10 | 21 | 0 | 4.35 | 0.934 | 0.073 | 1.405 | 0.201 |
| OP | 12 | 24 | 1 | 4.35 | 0.936 | 0.072 | 1.419 | 0.211 |
| OPTOF | 12 | 21 | 1 | 4.4 | 0.934 | 0.072 | 1.402 | 0.202 |

| | | | | | | | | |
|---|---|---|---|---|---|---|---|---|
| OP | 8 | 24 | 0 | 4.4 | 0.935 | 0.073 | 1.418 | 0.210 |
| OPTOF | 8 | 21 | 0 | 4.4 | 0.938 | 0.073 | 1.404 | 0.202 |
| OPTOF | 10 | 21 | 1 | 4.45 | 0.934 | 0.073 | 1.402 | 0.202 |
| OP | 10 | 24 | 1 | 4.45 | 0.935 | 0.072 | 1.418 | 0.211 |
| OP | 8 | 24 | 1 | 4.5 | 0.935 | 0.073 | 1.418 | 0.210 |
| OPTOF | 6 | 21 | 0 | 4.5 | 0.935 | 0.073 | 1.405 | 0.200 |
| OP | 6 | 24 | 0 | 4.5 | 0.937 | 0.071 | 1.419 | 0.211 |
| OPTOF | 8 | 21 | 1 | 4.5 | 0.938 | 0.074 | 1.401 | 0.200 |
| OPTOF | 4 | 21 | 0 | 4.55 | 0.935 | 0.071 | 1.400 | 0.202 |
| OPTOF | 6 | 21 | 1 | 4.6 | 0.935 | 0.072 | 1.401 | 0.201 |
| OP | 6 | 24 | 1 | 4.6 | 0.937 | 0.071 | 1.417 | 0.210 |
| OP | 4 | 24 | 0 | 4.6 | 0.939 | 0.072 | 1.417 | 0.211 |
| OPTOF | 4 | 21 | 1 | 4.65 | 0.935 | 0.071 | 1.400 | 0.202 |
| OP | 4 | 24 | 1 | 4.7 | 0.939 | 0.072 | 1.416 | 0.211 |
| OPTOF | 2 | 21 | 0 | 4.85 | 0.937 | 0.070 | 1.393 | 0.201 |
| OPTOF | 2 | 21 | 1 | 4.9 | 0.937 | 0.070 | 1.391 | 0.200 |
| OP | 2 | 24 | 0 | 4.95 | 0.950 | 0.069 | 1.415 | 0.208 |
| OP | 12 | 4 | 0 | 5.05 | 0.951 | 0.069 | 1.419 | 0.210 |
| OP | 2 | 2 | 1 | 5.1 | 0.950 | 0.068 | 1.417 | 0.209 |
| OP | 12 | 4 | 1 | 5.15 | 0.951 | 0.070 | 1.418 | 0.208 |
| OP | 10 | 4 | 0 | 5.2 | 0.955 | 0.069 | 1.418 | 0.208 |
| PSFTOF | 12 | 21 | 5 | 5.25 | 0.940 | 0.073 | 1.408 | 0.203 |
| PSFTOF | 10 | 21 | 5 | 5.35 | 0.938 | 0.072 | 1.406 | 0.201 |
| PSFTOF | 8 | 21 | 5 | 5.35 | 0.939 | 0.072 | 1.405 | 0.202 |
| OP | 10 | 4 | 1 | 5.35 | 0.955 | 0.068 | 1.416 | 0.209 |
| PSFTOF | 6 | 21 | 5 | 5.45 | 0.938 | 0.071 | 1.404 | 0.202 |
| OP | 8 | 4 | 0 | 5.5 | 0.962 | 0.071 | 1.416 | 0.203 |
| OP | 8 | 4 | 1 | 5.55 | 0.961 | 0.070 | 1.416 | 0.202 |
| PSFTOF | 4 | 21 | 5 | 5.65 | 0.939 | 0.069 | 1.399 | 0.203 |
| OP | 6 | 4 | 0 | 5.95 | 0.975 | 0.070 | 1.413 | 0.197 |
| OP | 6 | 4 | 1 | 6 | 0.975 | 0.070 | 1.413 | 0.196 |
| PSFTOF | 2 | 21 | 5 | 6.25 | 0.934 | 0.066 | 1.376 | 0.198 |
| OP | 4 | 4 | 0 | 6.8 | 0.999 | 0.073 | 1.405 | 0.180 |
| OP | 4 | 4 | 1 | 6.85 | 0.998 | 0.072 | 1.405 | 0.179 |
| OP | 8 | 24 | 5 | 7 | 0.942 | 0.065 | 1.388 | 0.199 |
| OP | 10 | 24 | 5 | 7 | 0.942 | 0.064 | 1.391 | 0.199 |
| OP | 12 | 24 | 5 | 7.05 | 0.943 | 0.064 | 1.390 | 0.199 |
| OP | 6 | 24 | 5 | 7.05 | 0.942 | 0.063 | 1.387 | 0.200 |
| OP | 4 | 24 | 5 | 7.1 | 0.942 | 0.063 | 1.383 | 0.199 |
| OPTOF | 12 | 21 | 5 | 7.15 | 0.942 | 0.065 | 1.373 | 0.190 |
| OPTOF | 6 | 21 | 5 | 7.2 | 0.940 | 0.065 | 1.371 | 0.189 |
| OPTOF | 10 | 21 | 5 | 7.2 | 0.941 | 0.065 | 1.371 | 0.188 |
| OPTOF | 8 | 21 | 5 | 7.2 | 0.945 | 0.067 | 1.372 | 0.188 |
| OPTOF | 4 | 21 | 5 | 7.25 | 0.939 | 0.064 | 1.365 | 0.186 |

| | | | | | | | | |
|---|---|---|---|---|---|---|---|---|
| OP | 2 | 24 | 5 | 7.5 | 0.948 | 0.060 | 1.376 | 0.191 |
| OPTOF | 2 | 21 | 5 | 7.55 | 0.938 | 0.062 | 1.357 | 0.184 |
| OP | 12 | 4 | 5 | 7.55 | 0.948 | 0.061 | 1.377 | 0.196 |
| OP | 10 | 4 | 5 | 7.7 | 0.953 | 0.061 | 1.380 | 0.192 |
| OP | 8 | 4 | 5 | 7.9 | 0.959 | 0.062 | 1.382 | 0.189 |
| OP | 6 | 4 | 5 | 8.3 | 0.973 | 0.064 | 1.380 | 0.182 |
| OP | 4 | 4 | 5 | 9.05 | 0.994 | 0.066 | 1.377 | 0.168 |
